# White matter variability, cognition, and disorders: a systematic review

**DOI:** 10.1101/2020.04.22.20075127

**Authors:** Stephanie J. Forkel, Patrick Friedrich, Michel Thiebaut de Schotten, Henrietta Howells

## Abstract

Inter-individual differences can inform treatment procedures and - if accounted for - have the potential to significantly improve patient outcomes. However, when studying brain anatomy, these inter-individual variations are commonly unaccounted for, despite reports of differences in gross anatomical features, cross-sectional and connectional anatomy. Brain connections are essential to facilitate functional organisation and, when severed, cause impairments or complete loss of function. Hence the study of cerebral white matter may be an ideal compromise to capture inter-individual variability in structure and function. We reviewed the wealth of studies that associate functions and clinical symptoms with individual tracts using diffusion tractography. Our systematic review indicates that tractography has proven to be a sensitive method in neurology, psychiatry, and healthy populations to identify variability and its functional correlates. However, the literature may be biased, as we determined that the most commonly studied tracts are not necessarily those with the highest sensitivity to cognitive functions and pathologies. Additionally, the hemisphere of the studied tract is often unreported, thus neglecting functional laterality and asymmetries. Finally, we demonstrate that tracts, as we define them, are not usually correlated with only one, but rather multiple cognitive domains or pathologies. While our systematic review identified some methodological caveats, it also suggests that tract-function correlations might be a promising biomarker for precision medicine. It characterises variations in brain anatomy, differences in functional organisation, and predicts resilience and recovery in patients.

## Introduction

If one stopped in a busy street and observed passers-by, one cannot help but observe that people are physically different. This diversity in appearance but also opinions, creativity, and morals has helped create a rich society. Individuals of different races, ethnicities, religious beliefs, socioeconomic status, language, and geographical origins make up our diverse community. Ever-evolving changes in our genome and adaptation to environmental factors have contributed to a range of genotypes (variability in genetic code) and phenotypes (variability in observable traits, *e*.*g*. eye colour) and the interaction between them (White & Rabargo-Smith, 2011). In the medical world, studying these inter-individual differences has led to the new disciplines of personalised and precision medicine. Inter-individual differences can help inform treatment procedures, and accounting for them has already improved patient outcomes and saved lives.

However, when turning to differences in brain anatomy, these inter-individual variations are relatively understudied (Glasser et al., 2016). It is often assumed that we share the same organisation of cognitive functions and underlying brain anatomy (Caramazza, 1986; Goldin *et al*., 2008; Greene *et al*., 2004; Johnson-Frey, 2004; Treu *et al*., 2020; Linden, 2020). For this reason, results from brain mapping studies are often depicted as group averages on template brains where inter-individual variability is considered an irrelevant deviation from the mean or considered to be a pathological change. In contrast, neuroanatomical studies often report or indeed study variability in individual brain structures (*e*.*g*. Sachs, 1892; Ono *et al*., 1990; Rademacher *et al*., 1993; Amunts *et al*., 1999, Caspers *et al*., 2006; Fornito *et al*., 2008), psychologists assume a Gaussian distribution of cognition and behaviour (Seghier & Price, 2018), and clinicians report differences in susceptibility to disorders and recovery (Forkel *et al*., 2014; Forkel *et al*., 2020). Although the existence of structural and functional variability is known, the ability to study interindividual variability across large populations and consider structural variability as having functional correlates has emerged only recently. This has been made possible through the availability of unique datasets with critical sample sizes and advances in computing power (Braver et al., 2010; Kanai & Rees, 2011; Dubois & Adolphs, 2016). The structure and function of the brain varies greatly between individuals and neuroimaging is sensitive to capture both sources of variability (Lerch et al., 2017; Gordon et al., 2017; Grasby et al., 2020; Tavor et al., 2016). On a structural level, measures of cortical surface area and thickness show hemispheric asymmetries that vary within the population (Kong et al., 2018). Brain morphology is also variable with half of the population having an additional gyrus, the paracingulate gyrus, in at least one hemisphere, for example (Fornito et al., 2008). Even primary cortical regions such as the motor, auditory, and visual cortices, are subject to anatomical variations (Uylings et al., 2005; Caulo et al., 2005; Leonard et al., 1998; Eichert et al., 2020) and associative cortical regions have variable cytoarchitectonic boundaries (Amunts et al., 1999). This body of literature indicates that a large amount of structural variability exists in primary cortical areas and associative cortices. Still, it is as yet unclear how observable behaviour and cognitive measures relate to these structural alterations.

There is increasing interest in understanding the brain’s structure-function relationship in the light of interindividual variability. Recent evidence has identified anatomical variations that are linked to differences in cognition and clinical outcomes (Forkel et al., 2020; Harrisson et al., 2020; Johnson et al., 2020; Taebi et al., 2020; Munsell et al., 2020; Wang et al., 2021). The neurosurgical literature is also increasing our understanding through mapping cognitive-anatomical variability in single case series during pre-, post, and intrasurgical imaging assessments (e.g. Vanderweyen et al., 2020). This multimodal brain mapping approach is able to reveal variation but also ‘atypical cases’, meaning the patients that do not fit expected assumptions of associations between one brain area or brain connection and a deficit in one cognitive domain. In the surgical setting, transcranial magnetic stimulation for presurgical planning (e.g. Giampiccolo et al., 2020; Mirchandani et al., 2020), deep brain stimulation (e.g. Calabrese, 2016; Akram et al., 2017), and direct electrical cortical stimulation during awake surgery (e.g. Puglisi et al., 2019; Middlebrooks et al., 2020) has been greatly aided by the consideration of inter-individual variability in white matter tracts estimated with tractography. However, there has not yet been a systematic attempt to capture this variability in connections across the entire brain and associate white matter phenotypes with cognitive profiles and clinical dimensions. It is, therefore, high time we included inter-individual variability and revisited the drawing board of neurology and psychiatry.

Clinical cases and mapping of inter-individual differences are beginning to explain the observed variance in cognitive and behavioural measures. As such, a better understanding of variability is crucial to explain differences in human abilities and disabilities and improve our clinical models and predictions (Seghier & Price, 2018). While the cerebral white matter may not be a functional agent *per se* (see Innocenti et al. 2017; Rockland, 2020), it constrains the brain’s functional organisation (Bouhali et al., 2014; Thiebaut de Schotten et al., 2017; Takemura & Thiebaut de Schotten, 2020) and leads to cognitive impairment or complete loss of function when severed (Geschwind et al. 1965a, 1965b). Hence, mapping white matter variability may be a useful surrogate measure to capture inter-individual differences in structure and function. Diffusion tractography has become an established non-invasive quantitative method to study connectional anatomy in the living human brain over the past 15 years (for reviews see Assaf et al., 2017; Jbabdi & Johanson-Berg, 2011). Tractography has been employed as a neuroimaging biomarker to link white matter phenotypes, meaning inter-individual variations in white matter networks, to cognition. These white matter phenotypes are a product of an environment-genotype interaction as has been demonstrated for the language and limbic networks (Su et al., 2020; Budisaljevic et al., 2016; Budisaljevic et al., 2015). Consequently, white matter networks are subject to variations over the lifespan and can change with training (Scholz et al. 2009; Lebel et al., 2019; Thiebaut de Schotten et al., 2014; Vanderauwera et al., 2018). Tractography has been shown to be highly sensitive in capturing these variations, which can be associated with inter-individual differences in neuropsychological measures in the healthy population (*e*.*g*. Catani et al., 2007, Thiebaut de Schotten et al. 2011; Howells et al. 2018) and clinical groups (*e*.*g*. Forkel et al., 2014; Forkel et al. 2020; Thompson 2017; Pacella et al. 2020; Alves et al. 2021). Therefore, tractography can be employed to study variability in the human brain and map functional white matter correlates.

Identifying consistent trends in the diffusion tractography literature may be a crucial step in mapping white matter phenotypes and their impact on cognition, hence a systematic review is timely. We focus here on studies that describe significant correlations between structural and continuous cognitive measures in healthy adults and psychiatric and neurological patients. For structure, we focus on volumetric or microstructural (e.g. fractional anisotropy, mean diffusivity) measures of white matter tracts that can be extracted from tractography reconstructions and voxel-wise measurements. We concentrate on neuropsychological tests in healthy volunteers and clinical scales in pathological populations to estimate cognitive-behavioural measures and clinical symptom severity. In this review, we summarise dimensional differences (i.e. correlations) between structural white matter connectivity (i.e. volumetric or microstructural) and cognition as a first step toward the systematic inclusion of inter-individual variability in neuroscience studies.

## Methods

We undertook a systematic review of published journal articles that correlated measures derived from white matter tractography with cognition or clinical symptoms, following PRISMA guidelines (Liberati et al., 2009).

The resources obtained from this study and created for this data are made available as supplementary material: https://github.com/StephForkel/PhenotypesReview.git

### Data Sources

A title/abstract search in MEDLINE and Scopus (which includes most of the EMBASE database, https://www.elsevier.com/solutions/embase-biomedical-research) was conducted. The search term ‘tractography’ returned a total of 5,303 in PubMed and 7,204 results in Scopus. We hence restricted our search (conducted on February 25th, 2020) to the following strings: (predictor OR “correlat*” OR regression OR “assoc*”) AND (tractography). Additional filters were applied to include only human adult studies published in English as final stage peer-reviewed articles in scientific journals. The search returned 1333 results on PubMed and 2380 results on Scopus, yielding 3,713 records. There were no internal duplicates within each database, and we excluded 1224 external duplicates between the databases. After removing duplicates from these lists, a total of 2489 results were screened.

### Data screening and eligibility

Figure 1 summarises the following workflow. During the screening, we applied further exclusion criteria leading to the exclusion of paediatric studies, non-human studies, pure methodological papers without behavioural correlates, correlations between tractography and physiological rather than behavioural measures (e.g. heart rate), non-brain studies (e.g. cranial nerves, spine, muscle), graph theory and tract-based spatial statistics (TBSS) studies, and case studies or mini-series (less than 10 participants/patients) and papers that reported no significant correlation. All studies reporting variability of white matter tracts using tractography that described a significant association with continuous cognitive measures, clinical symptom severity and/or continuous recovery were included. After screening of the abstracts, we retained a list of 466 references. Full text screening further identified studies that fulfilled the exclusion criteria defined above. This led to a total of 326 studies included in the final analysis.

**Figure 1.**
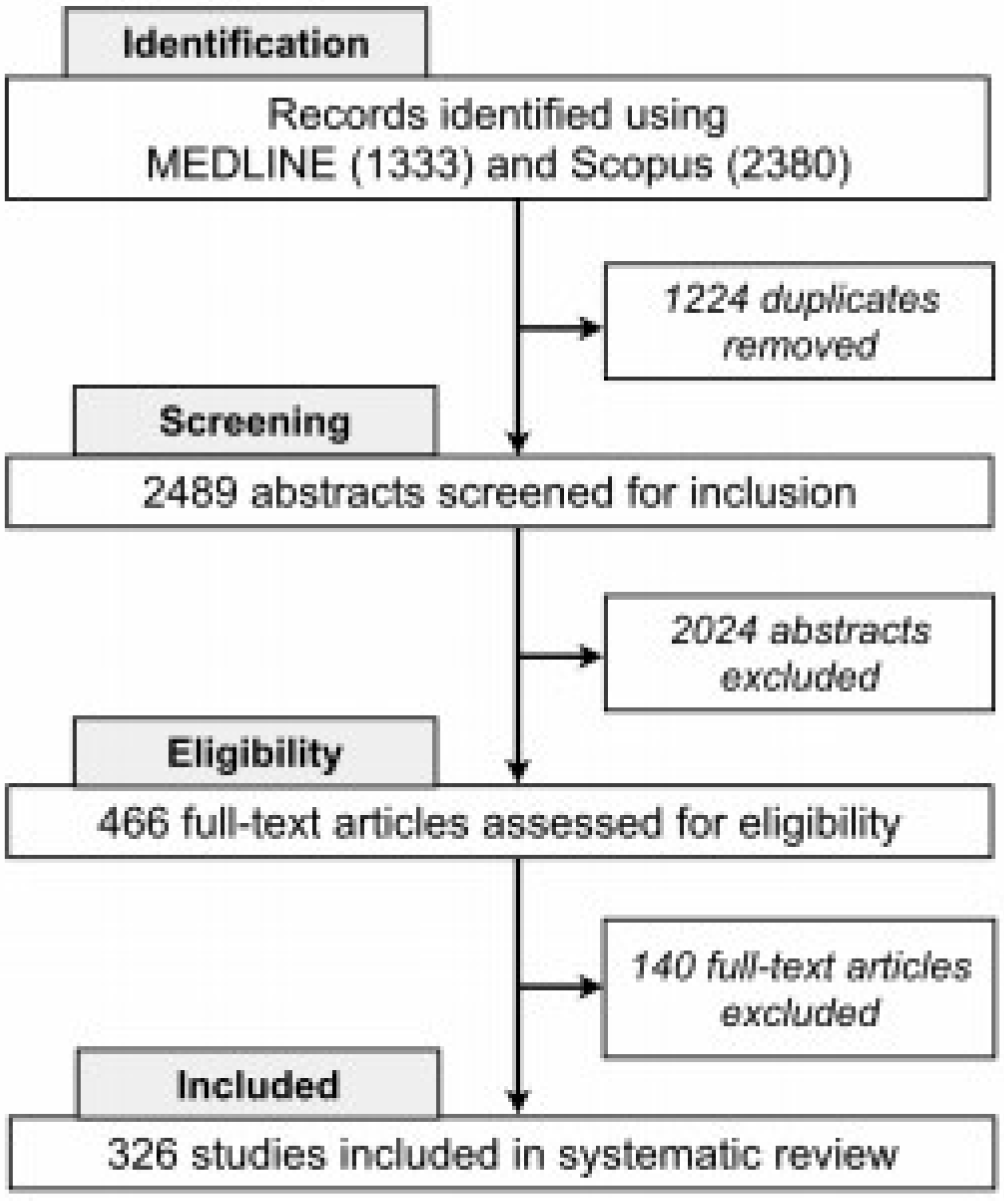
PRISMA flow chart

#### Study quality

The QUADAS quality assessment tool (Whiting et al., 2003) was adapted for the review to document the steps taken by each paper to avoid bias and justify and validate the protocols. The following criteria were used to rate publications: 1) Sufficient detail provided to reproduce the protocol, 2) clearly defined white matter tracts, 3) the groups, cognitive measures or clinical characteristics were reported.

### Data extraction

The following information was collected from the records: year of publication, group (e.g. healthy participants vs degeneration vs psychiatric vs neurological vs neurodevelopmental), sample sizes, left/right/unspecified hemisphere, tractography indices, label of white matter tracts, clinical symptoms, behaviour and/or cognitive domain, differential neuropsychological measures (*e*.*g*. Trail Making test), and finally the interaction between white matter tracts and neuropsychological assessments. The labelling of the groups (healthy participants vs degeneration vs psychiatric vs neurological vs neurodevelopmental) was aligned to current diagnostic criteria and categories of disease (e.g. DSM-5, IDC-11). The coding of the parameters is available from the supplementary material online (https://github.com/StephForkel/PhenotypesReview). As an example, a study in neurological patients measuring motor functions and the left corticospinal tract would be coded as: neurological*motor*CST_lh.

### Data synthesis and analyses

In the present synthesis of this dataset, we summarised degeneration, neurosurgical and common neurological symptoms as a neurological group. Similarly, the psychiatric group included adult neurodevelopmental and psychiatric studies. We also synthesised clinical symptoms, behaviour and/or cognitive domains using the following terms: We limited the cognitive domains taxonomy to the terms that are currently widely accepted in the literature, including attention, executive functions, language, memory, and reward. Terms defining behavioural domains included addiction, auditory, visual and motor behaviour, sleep, mood, social measures (*e*.*g*. theory of mind). To assess the sensitivity of tracts to clinical measures, we also included a “symptoms” dimension corresponding to neurological and psychiatric severity measures.

According to domains, the classification of the correlations was replicated three times by SJF, PF and HH and in case of disagreements, a consensus was reached. From these terms, we could extract variables of interest such as the number of correlations per tract (*i*.*e*. sensitivity or how likely a tract can correlate), but also the number of studies reporting significant correlations for that tract (*i*.*e*. popularity or how often studies report a significant correlation for that tract). A differentiation between sensitivity and popularity was made because many studies tested multiple associations between a tract and several cognitive measures. Therefore, if a study investigated the relationship between a tract and multiple functions, the sensitivity measure would increase with each significant correlation; however, the study would only be counted once for the popularity value. Subsequently, we also investigated the number of correlations reported according to the domains of interest mentioned above (*i*.*e*. specificity to cognitive domains: attention, executive functions, language, memory, and reward; behavioural domains: addiction, auditory, visual and motor behaviour, sleep, mood, social; and symptoms severity).

## Results

### The number of correlations and studies per tract

A total of 25 individual white matter tracts were reported to correlate with performance on neuropsychological tests and clinical symptoms (Figure 2). Amongst these, certain tracts were more commonly correlated with cognitive-behavioural measures (Figure 2A). We report here the number of studies that described correlations (Figure 2A) and the number of correlations per tract (Figure 2B). Showing this difference is essential, as some studies reported more than one tract correlation. Notably, commonly reported tracts (*i*.*e*. sensitivity) were not always those that were most systematically studied (*i*.*e*. popularity) indicated by the different number of studies per tract reported (Figure 2B).

**Figure 2.**
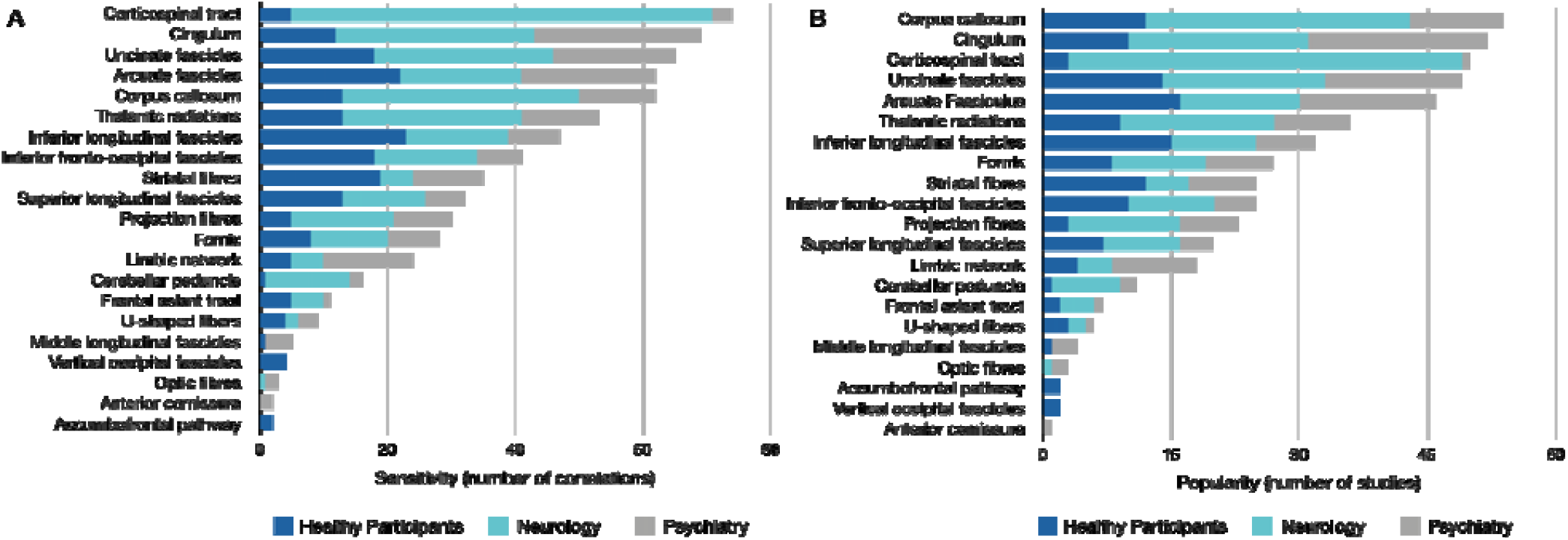
Frequencies of reported correlations (A) and the number of studies (B) per tract in each field of study (i.e. healthy participants, neurology, psychiatry). A high number of correlations indicates a high tract sensitivity, and the number of studies represents tract popularity.

Our review demonstrates that most results were reported for patients with neurological (45%) or psychiatric (29%) pathologies rather than controls (25%) (Figure 2). Additionally, the most studied tracts that have been reported to correlate with cognitive measures vary for each group. For example, most correlations reported in healthy participants were with the inferior longitudinal fasciculus, arcuate fasciculus, and striatal fibres (Figure 3A). In the neurological groups, correlations were mainly reported for the corticospinal tract, the corpus callosum, and the cingulum (Figure 3B). The cingulum, arcuate and uncinate fasciculus were the most prominent tracts to correlate with psychiatric symptoms (Figure 3C). The most correlated (i.e. sensitivity), however, does not mean the most commonly studied tracts (i.e. popularity) and might point towards a bias in the literature to focus on ‘target’ tracts rather than systematically studying the whole white matter. In healthy participants, the most ‘popular’ tracts were the arcuate, inferior longitudinal, and uncinate fasciculi (Figure 3D). For the neurological group, the most studied tracts were also the most sensitive tracts, namely the corticospinal tract, corpus callosum, and cingulum (Figure 3E). In the psychiatric group, the most sensitive and popular tracts were the cingulum, arcuate fasciculus and uncinate (Figure 3F).

**Figure 3.**
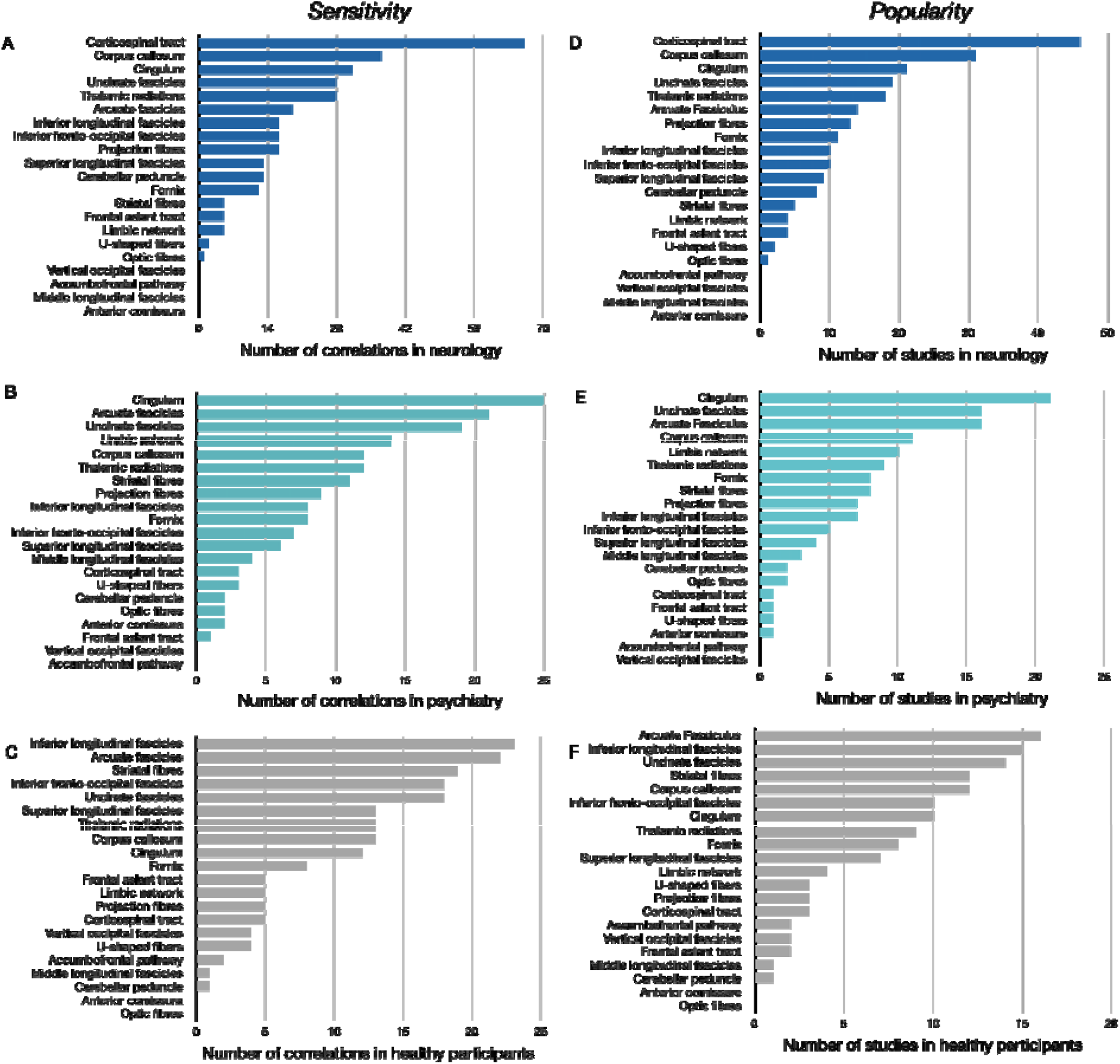
*“Tract bias” in the literature. Tract sensitivity (A-C) and tract popularity (D-F) in healthy participants, neurological group, and psychiatric group.* The number of correlations per tract is defined as ‘sensitivity’ or how likely a tract can correlate, whereas ‘popularity’ is defined as the number of studies reporting significant correlations for that tract. (Note: Data shown is the same as Figure 2 split by group)

### The number of correlations per domain

The analysis of correlations between tracts and cognitive domains showed no one-to-one correspondence between a white matter tract and a domain (Figure 4). The tracts that had the highest number of correlations (i.e. selectivity) with one domain were the corticospinal tract with the motor domain and the cingulum with executive functions (Figure 4). Additional figures showing all correlations per domain for all tracts are available in the supplementary material (https://github.com/StephForkel/PhenotypesReview.git). This summary also shows that the extent of a tract’s selectivity to one domain is often related to the diversity of the tract’s projections. For instance, the corpus callosum, which projects to most of the brain’s surface (Karolis et al., 2019) is associated domains, but the most common association for this pathway was with language measures. When separating the arcuate into its subdivisions (Catani 2005), this showed that its fronto-temporal segment was driving the domain specificity of the arcuate with language. In contrast, correlations with the anterior and posterior segments of the arcuate fasciculus were usually with aspects of the memory and attention domain (Figure 4).

**Figure 4.**
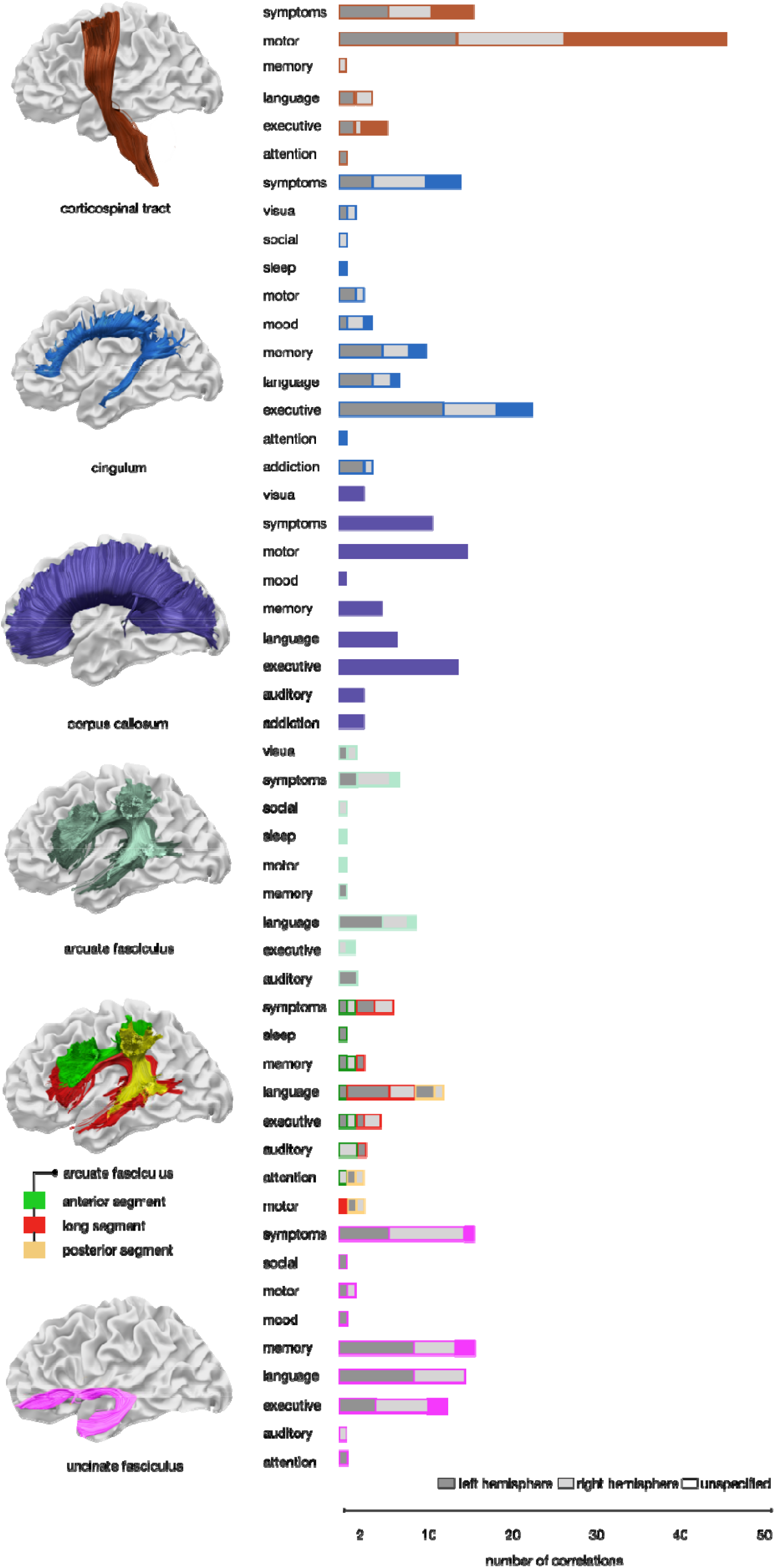
Specificity for tract x domain correlations. The number of correlations between cognitive, behavioural or clinical assessments and white matter tracts demonstrates that the concept of ‘one tract-one function’ does not hold. The figure shows the most studied tracts as identified by the current study. The other tract-domain correlations are available as supplementary figures (see supplementary figures).

### Hemispheric specialisation

Hemispheric specialisation is inconsistently reported in the current body of literature, with as many as 14% of studies not specifying if their correlations with a domain are for the left or right hemisphere tracts (Figure 5A). Amongst the 326 papers assessed, a total of 674 tract-function correlations with a p value reported of 0.05 or below were extracted. Within this data pool, an equal number of studies specified their results for the left (37.38%) and the right hemisphere (35.01%), while the remaining results (n=186) were unspecified (14%) or described commissural connections (19%) that cannot be attributed to either hemisphere. When looking at the distribution of significant correlations with cognitive measures it was evident that correlations with the left hemisphere are more commonly reported - or more commonly studied (Figure 5A). Specifying the hemisphere for commissural tracts is of course not required, however, this is essential when studying association or projection fibres, given the strong structural and functional lateralisation of the brain for some tracts and cognitive functions (e.g. Thiebaut de Schotten et al., 2011; Koralis et al., 2019).

**Figure 5.**
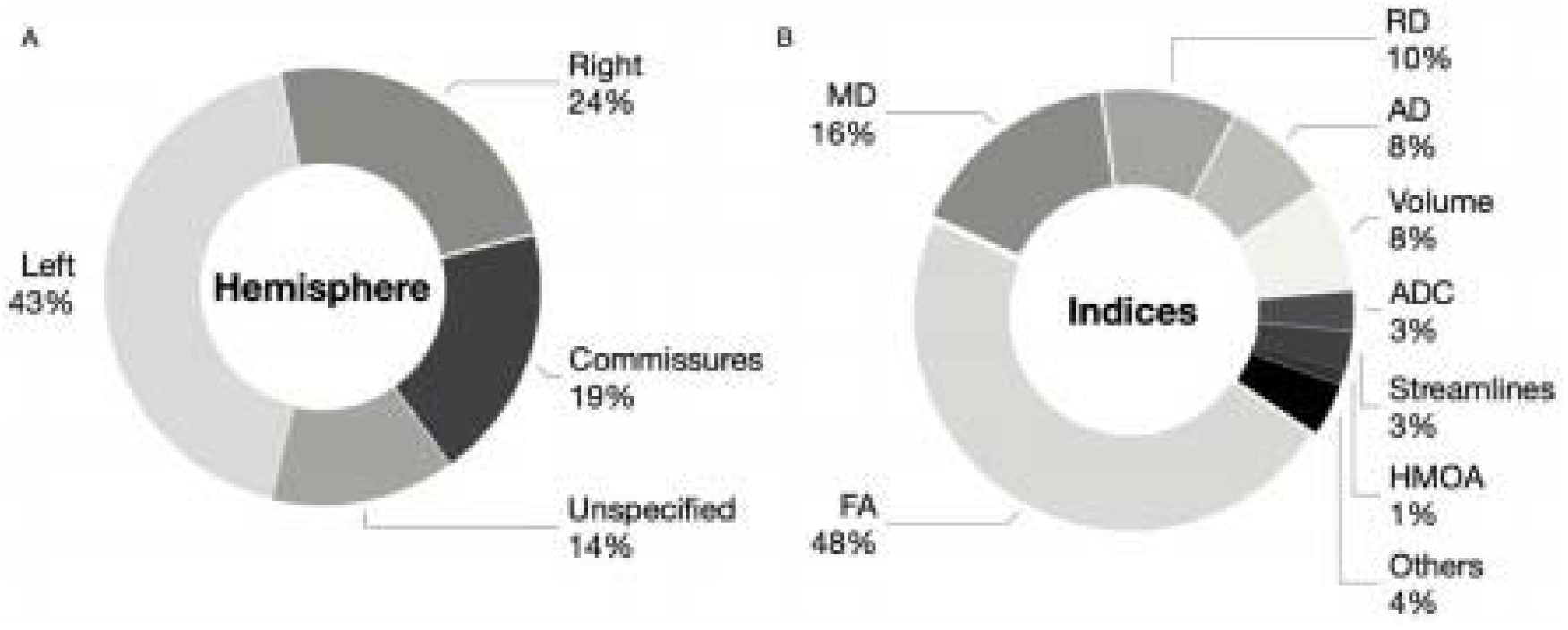
Summary of reporting of tract-domain correlations for each hemisphere of the brain (A) and diffusion indices (B). The results show that a greater number of correlations between tractography results in the left hemisphere and cognitive-behavioural measures exist in the literature and that studies with significant correlations most commonly use fractional anisotropy (FA). Mean diffusivity, MD; radial diffusivity, RD; axial diffusivity, AD; Apparent Diffusion Coefficient, ADC; Hindrance modulated orientational anisotropy, HMOA.

### Diffusion indices

A multitude of diffusion imaging methods have been developed and applied to the living brain. The first model to be widely applied to the study of the healthy living brain and pathological groups was diffusion tensor imaging (DTI). Using this model, various average properties of tissues within each voxel could be characterised by diffusion indices, including fractional anisotropy (FA), mean diffusivity (MD), number of streamlines, and voxels intersected by streamlines as a proxy of volume (mm^3^ or cm^3^). Volumetric measures accounted for 11% of the tract-domain correlations in the studied literature (Figure 5). These indices are computed using the diffusivity in each voxel, have been associated with microstructural properties and are used to indicate axonal damage or degeneration (Le Bihan, 1995; Pierpaoli & Basser, 1996; Beaulieu 2002; Ciccarelli et al., 2008; Afzali et al., 2021). Each index was extracted from the 326 studies and the results highlight that some measures are more commonly reported than others (Figure 5). Below we briefly discuss the meaning of these indices and their prevalence in the literature.

The quantitative apparent diffusion coefficient map (ADC) is a primary sequence in acute stroke imaging due to its sensitivity to early ischemic changes. The lesion cascade typically induces cytotoxic oedema which results in a quick drop in ADC by 30-50% (Moseley, 1990). ADC comprises about 3% of the correlational diffusion tractography literature. Another clinically useful surrogate measure of diffusion deficits is mean diffusivity (MD= λ1 + λ2 + λ3/3), a measure independent of tissue directionality and more prominently used in the literature at 16% (Figure 5). This map does not offer anatomical details but is sensitive to diffusion abnormalities, such as acute ischemic lesions (Lythgoe et al., 1997). The diffusivity measured along the principle axis (λ1) is referred to as axial diffusivity (also called longitudinal or parallel diffusivity as diffusivity is parallel to the axonal fibres, AD=λ1). While the diffusivity perpendicular to the fibres is calculated from the mean along the two perpendicular directions. This average of diffusivities along the two orthogonal axes (λ2, λ3) is denoted as perpendicular or radial diffusivity (RD=λ2 + λ3/2). Together, axial and radial indices make up 18% of the literature (Figure 5). The application of these measure is however still debated, as the direction and magnitude of the eigenvalue/eigenvector system relies on physical measures that are sensitive to noise and may be influenced by the estimated tensor ellipsoid and underlying pathologies (Wheeler-Kingshott & Cercignani 2009). As such, changes in axial diffusivity measurements, for example, could be related to intra-axonal composition, while radial diffusivity may be more sensitive to changes in membrane permeability and myelin density (Song et al., 2002). By far the most prominently used diffusion measure is fractional anisotropy (FA) which comprises 48% of the literature (Figure 5). The variance of all three eigenvectors (ν1-3) about their mean is normalised by the overall magnitude of the tensor and is referred to as fractional anisotropy (FA= 3((λ □ – λ)^2^ + (λ □ – λ) ^2^ + (λ □ – λ) ^2^) / 2 (λ □ ^2^ + λ □ ^2^ + λ □ ^2^)) (Jones 2009). The resulting FA index represents the fraction of the tensor that can be attributed to anisotropic diffusion (i.e. unequal diffusivity along directions) as the deviation from isotropy (i.e. equal diffusivity in all directions). FA designates free diffusivity (i.e. unhindered isotropic) with a value of 0, and constrained diffusion (i.e. anisotropic along one axis only) with the value of 1. Despite its wide adoption in the healthy and clinical literature, DTI and its voxel-wise measurements have their limitations (Dell’Aqua & Tournier, 2017; Meiher-Hein et al., 2017). Over the past years, advanced diffusion models have moved towards diffusion and fibre orientation density functions (fODF) to capture the complexity of white matter organisation using tract-specific measurements. One such measure is the apparent fibre density also referred to as hindrance modulated orientational anisotropy (HMOA) (Dell’Acqua et al., 2013; Dell’Aqua & Tournier, 2017), which currently comprise about 1% of the correlational diffusion tractography literature.

Together, these in vivo diffusion-based measurements allow connectional anatomy to be defined at different scales in health and disease. However, given the sensitivity of these diffusion indices to tissue characteristics and brain lesions (e.g. in the presence of oedema), they have to be interpreted carefully.

## Discussion

Over the course of the last fifteen years, there have been over 300 studies in human adults showing significant correlations between white matter tracts and cognitive measures. These correlations lend support to the concept of the importance of inter-individual differences in healthy participants and brain pathologies (e.g. neurological and psychiatric). Our systematic review of this literature demonstrates that tractography is a commonly used tool to study inter-individual variability and has proven to be a sensitive method in neurology, psychiatry, and healthy volunteers. Secondly, there may be a “tract bias” in the literature, as tracts that are commonly studied (high popularity) are not necessarily those that have the highest number of significant correlations (high specificity) for a given cognitive function or clinical symptoms. Finally, our review clearly shows that tracts, as we define them, are never correlated with only one cognitive domain or pathology.

Our investigation collated tract-function correlations across neurological, psychiatric, and healthy populations. Most tract-domain correlations in the literature were identified in studies of pathological groups rather than healthy participants (Figure 3). There are several possible explanations for this. This predominance may originate from the broader dispersion of data points associated with pathologies (i.e. more variability). As the presence of pathology causes higher variability in both anatomy and cognitive/clinical test scores, these variations are more likely to be detected by linear correlations. Another explanation for the high number of tract-domain correlations in clinical groups is that differences between healthy participants are often considered to be noise which can mean they are reduced during data processing (Kanai & Rees, 2011). While noise may contribute to the difference observed in controls, it is now clearly established that diffusion tractography can capture inter-individual differences that reflect some of the variations in the functioning of the brain (*e*.*g*. Powell et al., 2006; Vernooij et al., 2007; Lezari et al., preprint). An alternative hypothesis could be that current neuroimaging or cognitive and behavioural tests are not sensitive enough to disentangle noise from real variability in healthy participants systematically (Braver et al., 2010; Rousselet & Pernet, 2012). The latter may be improved by using finer-grained cognitive measures, higher resolution data, and better anatomical tract definitions.

Our review identified a total of 25 studied tracts that have been significantly correlated with cognitive measures in healthy participants or symptom severity in patients. The precise number of white matter tracts in the human brain remains unknown and variable estimates originate from the use of different methods. Most atlases suggest that around 26 tracts can be reliably identified with most tractography methods (Mori, 2005; Lawes et al., 2008; Catani & Thiebaut de Schotten, 2008; Mori et al., 2009; Thiebaut de Schotten et al., 2011; Rojkova et al., 2016). Some recent atlases further identify additional intralobar connections (Catani et al., 2012; Guevara et al., 2012; Catani et al., 2017; Guevara et al., 2020). This review reported on some additional tracts that have not yet been incorporated into atlases, including the accumbofrontal tract and the vertical occipital fasciculus (Martínez-Molina et al. 2019; Rigoard et al., 2011, Vergani et al., 2016; Yeatman et al., 2013; Vergani et al., 2014), while other tracts have not yet been widely used in the literature and therefore do not feature in this review (e.g. medial occipital longitudinal tract; Beyh et al. preprint). Our results also highlight a bias in the literature toward studying specific tracts that have very well-established functions (e.g. corticospinal tract) or are easy to dissect in clinical groups (e.g. cingulum). The omission of other tracts does, of course, not mean they are functionally irrelevant as shown by our sensitivity measure. For example, a high number of correlations were identified for the corticospinal tract and motor functions as can be expected. However, there were also some significant correlations with other tracts not typically associated with motor functions (e.g. arcuate fasciculus, uncinate fasciculus). It could thus be that some tract-function relationships are still poorly understood. Some may have non-linear or indirect relationships with function, for which correlational approaches are not appropriate. Further, understudied tracts may be more challenging to reconstruct due to limited anatomical guidelines or available algorithms (*e*.*g*. U-shaped fibres, Attar et al., 2020, Mandelstam, 2012; Maffei et al., 2019a; 2019b).

For the most sensitive, or commonly correlated, tracts, several functions were reported. Our results show that even the corticospinal tract that is primarily studied within the motor domain (62.21% of correlations, Figure 4), showed a non-uniform functional profile. For instance, some studies reported correlations between the corticospinal tract and executive functions (8.11%) and language/speech processes (5.4%). For other tracts, the correlations were even more diverse. For example, the cingulum correlated with psychiatric symptom severity (20.29%), memory (14.49%) and language measures (10.14%). Such results support the idea of hierarchical brain organisation with some tracts involved in mediating many functions, whereas others may have a more specific functional profile (Pandya and Yeterian 1990). While the number of associations is very likely to be biased by several factors including prior hypotheses that a given tract is involved in a specific function, a recent study mapped a total of 590 cognitive functions, as defined by a meta-analysis of activations derived from fMRI paradigms, onto a white matter atlas (Thiebaut de Schotten et al., 2020). This first functional atlas of the white matter demonstrated that one tract can be relevant for multiple functions. Another possible interpretation of this finding is that human-ascribed definitions of white matter tracts are too coarse to be specific to only one given function. For example, segmenting the arcuate fasciculus into three components in line with the early work (Catani et al., 2005) shows correlations with more domain specificity than correlations with the entire arcuate fasciculus. This may call for finer-grained white matter divisions or data-driven approaches to identify segments of white matter that may be related to specific functions (see, for example, Foulon et al., 2018; Nozais et al., in press).

We also show differential patterns between healthy participants and pathological groups. One such example is the size of the uncinate fasciculus that has been primarily associated with memory in healthy ageing (Sasson et al., 2013), with psychopathy in psychiatric studies (*e*.*g*. Craig et al., 2009), and language in neurological studies (*e*.*g*. D’Anna et al., 2016). Similarly, the size of the arcuate fasciculus has been implicated in learning new words in healthy participants (Lopez-Barroso et al., 2013), which supports the role of the arcuate fasciculus as the mediator between the temporal-parietal-frontal cortices and the neural substrate for the phonological loop (Baddley et al., 1998; Catani et al., 2005; Baddley 2007; Buchsbaum & D’Eposito, 2008; see Baddley & Hitch, 2019 for a recent review on phonological loop). Recently this hypothesis was supported by intraoperative direct cortical stimulation in neurosurgical patients (Duffau et al., 2008; Papagano et al., 2017). In psychiatric and neurological patients, damage to the arcuate fasciculus was associated with auditory hallucinations in schizophrenia (Catani et al., 2011), aphasia severity in stroke (Forkel et al., 2014), and repetition deficits in primary progressive aphasia patients (Forkel et al., 2020). Therefore, the functions associated with a tract might not purely be a product of the cortical regions the white matter connects to but instead rely on the interplay of one region with another. When pathology is introduced into this delicate network, differential patterns of symptoms may reflect the variable impact on brain regions within such a network. Furthermore, the pathophysiological mechanisms are different across pathologies and have different long-range effects on connected regions (e.g. Catani et al., 2005).

There are limitations associated with tractography that may have influenced the studies summarised in the current work. We set out to systematically review tract-function correlations irrespective of these limitations, to identify broad patterns, however, it is essential to caveat this study by stating what tractography can and cannot do when interpreting results. While tractography has proven useful for research and clinical applications, interpretation of voxel-based indices presents challenges (Dell’Acqua & Tournier, 2019). When considering the resolution of diffusion data, for example, diffusion indices are averaged across and within voxels, which may mask meaningful changes. For research purposes, the voxel size is typically 2*2*2mm, while the voxel sizes are often larger for clinical acquisitions leading to even lower spatial resolution. As such, a research acquisition with an 8mm^3^ voxel is likely to contain an inhomogeneous sample of tissue classes, intra- and extracellular space, and axons of different densities and diameters. This multi tissue composition within voxels can pose challenges for the study of projection and commissural fibres and afflict tractography reconstructions with false positive and negative reconstructions. A recent preprint also looked at the commonly reported index of FA and demonstrated that multimodal approaches can help detect white matter behaviour relationships that are not detected with FA alone (Lezari et al., preprint). This study also raised the notion of the need for sufficiently powered samples to detect changes in myelin in relation to behaviour.

The diffusion signal itself is also inhomogeneous across the brain. As a result, areas such as the orbitofrontal cortex and anterior temporal cortex are often distorted. Methodological advances partially correct for these distortions (e.g. TOPUP, Andersson et al., 2003) and disentangle some of these components to reconstruct crossing fibres and extract tract-specific measurements (see Dell’Acqua & Tournier, 2019). However, most studies included in this review used diffusion tensor algorithms rather than advanced algorithms and indices (see Figure 5, HMOA 1% of studies). While recent research studies have the methodological means to mitigate such distortions (*e*.*g*. Andersson et al., 2003), most current clinical studies still suffer from these limitations, potentially explaining the lack of tract domain specificity.

Another source of inconsistencies originates from incoherent reporting of the anatomy. For example, many studies did not specify which hemisphere was studied or collapsed their white matter across both hemispheres and correlated the averaged anatomy with cognitive and behavioural measures. Collapsing measurements from anatomical features across both hemispheres might prove problematic for white matter tracts that are subject to more considerable inter-individual variability and subsequently might get over- and underrepresented in each hemisphere (*e*.*g*. Catani et al., 2007; Thiebaut de Schotten et al., 2011; Rojkova et al. 2016, Croxson et al., 2018; Howells et al. 2018; Howells et al. 2020). Further, while the concept of a strict hemispheric dichotomy might be seen as overly simplistic (*e*.*g*. Vingerhoets, 2019), splitting the measurements by hemisphere may reveal useful insights and higher specificity into the contribution of either side to a measured cognitive behaviour or disorder (Floris & Howells, 2018). Another limitation comes from inconsistencies in the classification of white matter tracts. For instance, the superior longitudinal fasciculus (SLF) was often considered in its entirety without specifying which branch was studied. When branches were specified, a variety of terminologies were used, including the three branches (SLFI-III, Thiebaut de Schotten et al., 2011), or a lobar-based segmentation into a SLFtp (temporal projections) and SLFpt (parietal projections) (*e*.*g*. Nakajima et al. 2019). Another example is the arcuate fasciculus that was sometime considered in its entirety and sometimes split into several branches (*e*.*g*. Catani et al., 2005; Kaplan et al., 2010). Perhaps due to early anatomical descriptions where the terminology was used interchangeably and incorrectly, we are still faced with a body of literature that uses the terms SLF and arcuate interchangeably. This confusion may have come about in the literature, as the SLF system was not easily dissected in the human brain using either post-mortem methods or tractography due to crossing fibres. In fact, this fronto-parietal network was first described in the monkey brain and only subsequently identified in the human brain using diffusion imaging (Makris et al. 2005) and diffusion tractography (Thiebaut de Schotten et al., 2011). While there is some overlap between both networks, such as the SLF-III and the anterior segment of the arcuate fasciculus, the other branches and segments are distinct. From an anatomical and etymological perspective, the superior longitudinal fasciculus should be ascribed solely fronto-parietal connections (*i*.*e*. “superior and longitudinal”; Thiebaut de Schotten et al., 2011) whereas the arcuate fasciculus should be considered to be the fronto-temporal connection (i.e. ‘arching’ around the Sylvian fissure; Catani et al., 2005). Recent attempts have synthesised this literature, suggesting using the term superior longitudinal system (SLS) to include the arcuate fasciculus stricto sensu and the three branches of the SLFs in one multilobar fibre system (Mandonnet et al., 2018; Vavassori et al., 2021). Another controversy in the literature is the differentiation between the posterior segment of the arcuate fasciculus and the vertical occipital fasciculus (Martino & Garcia-Porrero, 2013; Bartsch et al., 2013; Bullock et al., 2019; Weiner et al., 2017). The anatomy of the VOF has been verified using tractography (Yeatman et al., 2014; Keser et al., 2016; Briggs et al., 2018; Schurr et al., 2019; Panesar et al., 2019), postmortem dissections (Vergani et al., 2014; Gungor et al., 2017; Palejwala et al., 2020), and comparative anatomy (Takemura et al., 2017). These descriptions were scrutinised against historical postmortem descriptions from Wernicke (Yeatman et al., 2014), Sachs (Vergani et al., 2014) and the Dejerine’s (Bugain et al., 2021). Anatomically, the vertical occipital fibre system and the posterior arcuate segment are distinct bundles in terms of their trajectories and cortical terminations. More precisely, the vertical occipital fasciculus projections are in the occipital lobe and the posterior segment of the arcuate fasciculus projections are in the posterior temporal and parietal lobes (Weiner et al., 2017). Another example is the differential and synonymous use of the terminologies external capsule (Rilling et al., 2012), external/extreme fibre complex (Mars et al., 2016), inferior fronto-occipital fasciculus (Forkel et al., 2014; Sarubbo et al., 2015), and inferior occipitofrontal fasciculus (Kier et al., 2004). The difference in terminology is owed mainly to the description of these tracts using different methods (Forkel et al., 2014) and some consensus is certainly needed to improve consistency in the literature (Maier-Hein et al., 2017; Mandonnet et al., 2018; Vavassori et al., 2021). Another tract that appears under two terminologies in the literature is the medial occipital longitudinal tract (MOLT) relevant for visuospatial processing (Beyh et al., preprint). This tract has previously been referred to as the ‘sledge runner’ (Vergani et al., 2014).

Additionally, to harvest the interindividual variability results, this review focused on continuous cognitive and clinical measures obtained from correlations to associate them with white matter phenotypes. As such, we did not separate distinct structural subtypes (*e*.*g*. Ferreiera et al. 2020; Forkel et al., 2020) and did not take different diffusion matrices (*e*.*g*. fractional anisotropy vs mean diffusivity) or tractography algorithms (*e*.*g*. tensor vs HARDI) into account. Some of these parameters may be more sensitive and specific than others as discussed above.

However, some measures were underrepresented in our systematic review which prevented any valid comparison (Figure 5). Finally, while correlational research indicates that there may be a relationship between two variables (*e*.*g*. structure and function), it cannot assume causality and prove that one variable causes a change in another variable (e.g. Rousselet & Pernet, 2012). This means that from the correlation data reviewed in this study, it is impossible to determine whether anatomical variability is driving behaviour or if the anatomy results from an expressed behaviour (*i*.*e*. directionality problem). In addition, it is also not possible to know whether a third factor mediates the changes in both variables and that the two variables are in fact not related (*i*.*e*. third variable problem). Future studies using correlational tractography may benefit from exploring other statistical frameworks such as Bayesian methods to get closer to establishing causal relationships between variables (Pacella et al., 2019). Systematic reviews cannot answer all clinically relevant questions as they are retrospective research projects and as such subject to bias. One bias that was not able to mitigate in this review was the positive publication bias. We did not report negative correlations as they are not coherently reported in the literature. This review aimed to systematically evaluate and summarise current knowledge and going forward new initiatives such as registered reports should help reduce the positive publication bias associated with the clinical-anatomical correlation method (for an example see Lazari et al., preprint). Functional white matter atlases (Thiebaut de Schotten et al., 2020) can also help to decode cognitive networks and individual assessment battery correlations (Forkel et al. preprint; Dulyan et al., preprint; Talozzi et al., preprint).

In conclusion, acknowledging and objectively quantifying the degree of variability between each of us, particularly when it comes to brain anatomy, will potentially have a far-reaching impact on clinical practice. While some methodological refinement is needed in the field of white matter tractography (e.g. Wasserthal et al., 2018; Maier-Hein et al., 2017; Grisot et al. 2021), preliminary evidence indicates that differences in white matter phenotypes are beginning to explain disease progression and differential symptom presentations (Forkel et al., 2020). Variability in structural brain connections can also shed light on why current invasive and non-invasive treatments and therapies help some but not all patients (Lunven et al., 2019; Parlatini et al., under review; Sanefuji et al., 2017). These findings are encouraging, as we move towards more personalised approaches to medicine. With the improvements suggested in this systematic review, tract-function correlations could be a useful adjunct in studies predicting resilience and recovery in patients.

## Data Availability

All data is available online.

https://github.com/StephForkel/PhenotypesReview.git

## Acknowledgements

The authors would like to thank our reviewers and the Groupe d’imagerie neurofonctionnelle (GIN) whiteboard team for helpful discussions.

## Supplementary material

https://github.com/StephForkel/PhenotypesReview.git

